# Epidemiology of Hepatitis B Virus Co-infection Among People Living with HIV in Four Countries in Sub-Saharan Africa

**DOI:** 10.1101/2025.11.16.25340356

**Authors:** Josphat Kosgei, Walter Jaoko, Julius Oyugi, Natalie Burns, Jonah Maswai, John Owuoth, Valentine Sing’oei, Emmanuel Bahemana, Abdulwasiu B. Tiamiyu, Hannah Kibuuka, Leigh Anne Eller, Julie A. Ake, Neha Shah, Trevor A. Crowell, the African Cohort Study (AFRICOS) Study Group

## Abstract

**Background:** Hepatitis B virus (HBV) and human immunodeficiency virus (HIV) share transmission routes and are associated with morbidity in sub-Saharan Africa. Understanding the prevalence and demographic predictors of HBV co-infection among people living with HIV (PLWH) can inform integrated screening and treatment strategies.

**Methods:** We analyzed enrollment data from the African Cohort Study (AFRICOS), a prospective study of participants aged ≥15 years with and without HIV across 12 clinical sites in Kenya, Uganda, Tanzania, and Nigeria. HBV was diagnosed via hepatitis B surface antigen (Bio-Rad, Hercules, CA). Descriptive statistics summarized HBV prevalence by HIV status and demographic characteristics. Relative risk (RR) of HBV among PLWH compared to people without HIV was calculated. Among PLWH, multivariable logistic regression identified odds ratios (ORs) and 95% confidence intervals (CIs) for factors independently associated with HBV co-infection.

**Results:** Among 4,221 participants with valid HBV results, 3,433 were PLWH. HBV prevalence was 4.7% among PLWH and 4.3% among people without HIV (RR: 1.09; 95% CI: 0.76-1.56). Among PLWH, HBV prevalence was highest in Nigeria and lowest in Kenya (11.1% vs. 2.6%, p<0.001), higher among males than females (6.8% vs. 3.8%, p<0.001, and highest in participants aged ≥40 years and lowest in participants aged 15–19 years (5.8% vs. 0.2%, p<0.001). In multivariable analysis, male sex (OR: 2.28 [95%CI: 1.63-3.18]), Nigeria residence (OR: 5.19 [95%CI: 3.35-8.04]), and age ≥40 (OR: 2.90 [95%CI: 1.69-4.99]) were independently associated with HBV co-infection.

**Conclusion:** Routine HBV screening and targeted interventions are needed within HIV programs, especially for older males and in high-prevalence settings like Nigeria.

## Introduction

Hepatitis B virus (HBV) and human immunodeficiency virus (HIV) are two major global health concerns that frequently co-occur due to shared transmission routes, including condomless sexual contact, perinatal exposure, and parenteral routes such as injection drug use and unsafe medical practices. Co-infection with HBV and HIV presents significant clinical challenges, as it is associated with more rapid progression of liver disease, higher rates of chronic HBV infection, and increased liver-related morbidity and mortality compared to mono-infection with either virus^1^. Beyond liver disease, HBV co-infection among PLWH has also been associated with higher rates of hospitalization due to non-AIDS-defining infections, including bacterial and non-specific inflammatory conditions, underscoring the broader systemic impact of co-infection^2^.

Sub-Saharan Africa bears a substantial burden of both HIV and HBV, with overlapping endemicity in many countries^3^. Although antiretroviral therapy (ART) regimens that are active against both viruses have improved the prognosis of co-infected individuals, HBV remains a major contributor to liver-related complications among people living with HIV (PLWH), particularly in resource-limited settings where HBV screening and vaccination coverage remain low^4^.

Understanding the epidemiologic characteristics of HBV among PLWH is critical for informing integrated prevention, diagnosis, and treatment strategies. Previous studies have reported variable prevalence of HBV among PLWH across Sub-Saharan Africa, with co-infection influenced by geographic location, age, sex, and behavioral or healthcare-related exposures^5^. However, large-scale comparative studies evaluating HBV prevalence in people with and without HIV in this region remain limited.

We assessed the prevalence of HBV among a large cohort of individuals from four Sub-Saharan African countries—Kenya, Uganda, Tanzania, and Nigeria—with a particular focus on PLWH. We further examined demographic and regional associations of HBV co-infection among PLWH using multivariable logistic regression. Our findings contribute to the growing evidence base needed to guide public health policies and clinical management strategies for HIV/HBV co-infected populations in Sub-Saharan Africa.

## Methods

We analyzed prevalence and socio-demographic factors associated with hepatitis B co-infection among PLWH in the African Cohort Study (AFRICOS). AFRICOS is an open-ended prospective cohort study, that enrolled adults and adolescents aged 15 years and above, with and without HIV, at clinical sites in Kenya, Tanzania, Uganda, and Nigeria that are supported by the U.S. President’s Emergency Plan for AIDS Relief (PEPFAR) via the U.S. Military HIV Research Program (MHRP).

HBV was diagnosed by the presence of hepatitis B surface antigen detected using an enzyme-linked immunosorbent assay (ELISA) (Bio-Rad, Hercules, CA). Participants with a valid HBV test at any study visit were included in the analytic population using data from the time of their first valid HBV test. Demographic variables included age, sex, and country. Participants were categorized into age groups (15-19 years; 20-24 years; 25-39 years; 40 years and above).

Descriptive statistics were used to summarize HBV prevalence by HIV status and to characterize the demographic distribution of the analytic population. Categorical variables, including HIV and HBV status, sex, country of origin, and age group, were summarized using frequencies and percentages. HBV prevalence was calculated as the proportion of individuals testing positive for HBV among participants with a valid test, stratified by HIV status.

HBV prevalence was compared between people with and without HIV using a chi-square test, and a relative risk (RR) was calculated with 95% Wald confidence interval (CI).

Among PLWH, prevalence of HBV co-infection by age group, sex, and country were compared using chi-square tests or Fisher’s exact tests (for variables with any cell size <5). A p-value of <0.05 was considered statistically significant.

To identify factors independently associated with HBV co-infection among PLWH, a multivariable logistic regression model was constructed. The outcome variable was HBV co-infection (present vs. absent), and independent variables were decided a priori to include sex, country, and age group. Unadjusted and adjusted odds ratios (ORs) with 95% CIs were reported for each independent variable, along with associated p-values. All statistical analyses were conducted using R version 4.3.2^6^.

The AFRICOS study received ethical approval from Institutional Review Boards in Kenya, Uganda, Tanzania, Nigeria, Walter Reed Army Institute of Research, and all collaborating institutions. All participants, including both adults and minors, provided written informed consent or assent prior to enrollment. For minors, assent was obtained alongside written consent from their parents or legal guardians. Participant recruitment commenced on 21 January 2013 and remains ongoing.

## Results

### Participant Characteristics and HBV Prevalence

Among 4,270 participants enrolled between January 2013 and October 2024, 4,221 (98.9%) had a valid HBV test at any visit, of whom 3,433 (81.3%) were living with HIV and 788 (18.7%) were not living with HIV. A total of 195 (4.6%) were diagnosed with HBV at the visit with their first valid test. The prevalence of HBV among PLWH was 4.7% (161/3433) while it was 4.3% (34/788) among people without HIV, corresponding to a relative risk of 1.09 (95% CI: 0.76-1.56, p=0.651).

### Hepatitis B Co-Infection Among People Living with HIV

Of the 3,433 PLWH, 615 (17.9%) were from Uganda, 658 (19.2%) from Tanzania, 389 (11.3%) from Nigeria, and 1,771 (51.6%) from Kenya (Table 1). Most PLWH in our study were female (58.0%) and most were aged 25 years and above (39.6% aged 25-39 and 37.5% aged ≥40). The prevalence of HBV co-infection among PLWH varied by country, with Nigeria having the highest prevalence (11.1%) and Kenya having the lowest (2.6%, p<0.001; Figure 1A). The prevalence of HIV/HBV co-infection was higher among males (6.8%) as compared to females (3.2%, p<0.001; Figure 1B). The prevalence of HIV/HBV co-infection also increased with age, from 0.2% among PLWH aged 15-19 years to 5.8% among PLWH aged 40 years or older (p<0.001; Figure 1C).

**Table 1:** Demographic Distribution of Hepatitis B Co-infection Among People with HIV(n *=* 3433)

|  | Overall<br>(N=3433) | HBV+<br>(N=161) | HBV-<br>(N=3272) | p-<br>value<sup>1</sup> | Unadjusted OR (95%<br>CI) | p-value | Adjusted OR (95%<br>CI) | p-value |
| --- | --- | --- | --- | --- | --- | --- | --- | --- |
|  | n (col%) | n (row%) | n (row%) |  |  |  |  |  |
| **Country** |  |  |  | <0.001 |  |  |  |  |
| Uganda | 615 (17.9%) | 40 (6.5%) | 575 (93.5%) |  | 2.61 (1.69, 4.03) | <0.001 | 2.80 (1.81, 4.33) | <0.001 |
| Kenya | 1771 (51.6%) | 46 (2.6%) | 1725 (97.4%) |  | Reference |  |  |  |
| Tanzania | 658 (19.2%) | 32 (4.9%) | 626 (95.1%) |  | 1.92 (1.21, 3.05) | 0.005 | 2.10 (1.32, 3.34) | 0.002 |
| Nigeria | 389 (11.3%) | 43 (11.1%) | 346 (88.9%) |  | 4.70 (3.05, 7.24) | <0.001 | 5.19 (3.35, 8.04) | <0.001 |
| **Sex** |  |  |  | <0.001 |  |  |  |  |
| Male | 1441 (42.0%) | 98 (6.8%) | 1343 (93.2%) |  | 2.23 (1.61, 3.08) | <0.001 | 2.28 (1.63, 3.18) | <0.001 |
| Female | 1987 (58.0%) | 63 (3.2%) | 1924 (96.8%) |  | Reference |  |  |  |
| Missing | 5 | 0 | 5 |  |  |  |  |  |
| **Age**<br>**(years)** |  |  |  | <0.001 |  |  |  |  |
| 15-19 | 403 (11.8%) | 1 (0.2%) | 402 (99.8%) |  | Reference (combined 15-24 years category) |  |  |  |
| 20-24 | 382 (11.1%) | 16 (4.2%) | 366 (95.8%) |  |  |  |  |  |
| 25-39 | 1357 (39.6%) | 69 (5.1%) | 1288 (94.9%) |  | 2.42 (1.41, 4.15) | 0.001 | 2.70 (1.57, 4.65) | <0.001 |
| ≥40 | 1286 (37.5%) | 75 (5.8%) | 1211 (94.2%) |  | 2.80 (1.64, 4.77) | <0.001 | 2.90 (1.69, 4.99) | <0.001 |
| Missing | 5 | 0 | 5 |  |  |  |  |  |

<sup>1</sup>p-value from chi-squared test or Fisher's exact test (for variables with any cell size <5). Missing values were excluded from these tests.

**Figure 1.**
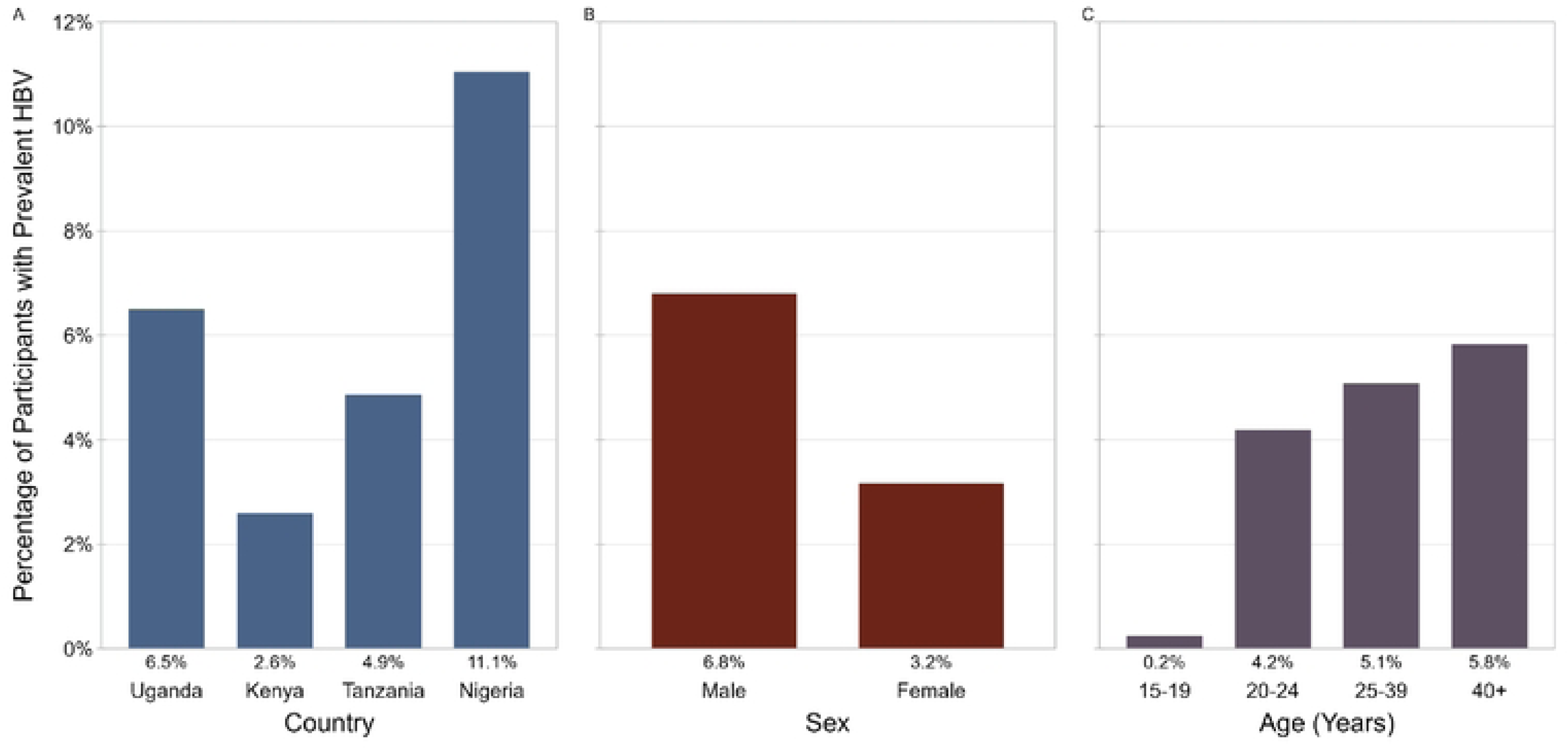
Prevalence of Hepatitis B among people with HIV by country, sex, and age group.

### Factors Associated with HBV Co-Infection among People Living with HIV

Logistic regression was applied to identify factors independently associated with HBV co-infection among PLWH (Table 1). Males were noted to be at over twice the odds of HBV co-infection compared to females (OR [95% CI]: 2.28 [1.63-3.18]; p<0.001). Participants from Nigeria had about five times higher odds (5.19 [3.35-8.04]; p<0.001) compared to participants from Kenya, with the older age groups having significantly higher odds of co-infection (25-39 years: 2.70 [1.57-4.65]; p<0.001; ≥40 years: 2.90 [1.69-4.99]; p<0.001) compared to participants aged 15-24 years (Figure 2).

**Figure 2.**
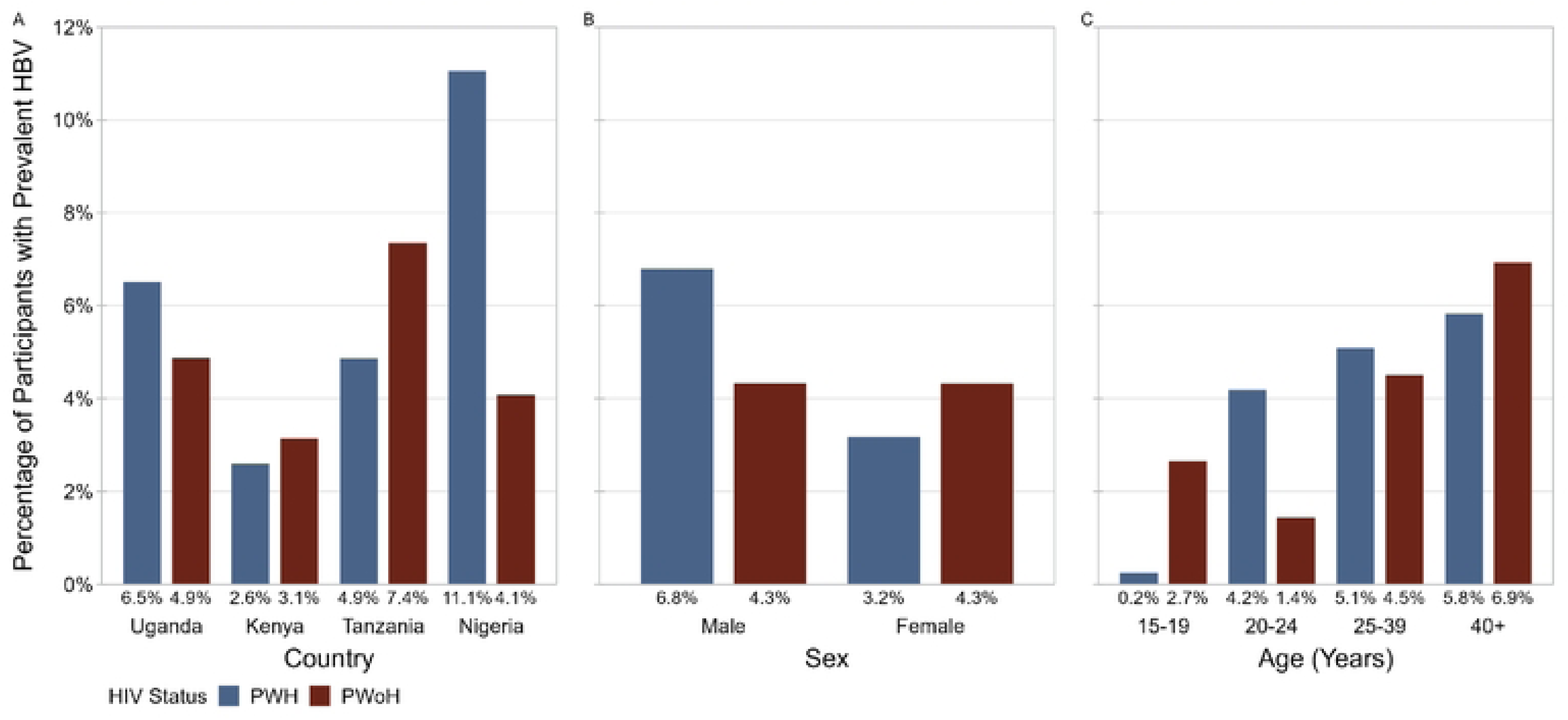
Prevalence of Hepatitis B by HIV status and by country, sex, and age group.

## Discussion

The overall HBV prevalence of 4.7% among PLWH in our cohort is slightly lower but broadly consistent with regional estimates from prior studies, which have reported co-infection rates ranging from 5% to 15% across Sub-Saharan Africa, depending on the population and diagnostic methods used^5,7,9^.

Nigeria exhibited the highest co-infection prevalence (11.1%), aligning with previous research that has identified West Africa as a region with particularly high HBV endemicity^8^. In contrast, Kenya had the lowest prevalence (2.6%), consistent with prior studies indicating a lower HBV burden in East Africa^9^. This difference observed across countries likely reflects differences in regional HBV endemicity, transmission patterns, and health system factors. West Africa has historically high HBV endemicity, partly due to more common perinatal and early childhood transmission, whereas East Africa tends to have lower chronic HBV rates with more horizontal transmission later in life^8,9^. Other factors may include variability in HBV vaccination coverage, as well as differences in diagnostic practices, healthcare access, and population-level risk factors. These factors influence the burden of HBV co-infection among PLWH in different settings. Although slightly lower, our findings are consistent with studies from other African regions. Shivakumar et al. (2024) reported an HBV prevalence of 7.1% among adults living with HIV in South Africa, a region that bears a significant burden of the HIV epidemic^10^. Similarly, a systematic review from Ethiopia estimated HBV prevalence among PLWH to range from 5% to 17%^11^, while a study in Senegal reported an HBV seroprevalence of 11.9% among PLWH^12^. In Europe, the EuroSIDA study found a 9% HIV/HBV co-infection prevalence, highlighting the consistent global intersection of these infections^13^. Additionally, the meta-analysis by Tengan et al., reported a pooled HBV prevalence of 7.0% among PLWH in Latin America and the Caribbean, suggesting that HBV co-infection remains a significant challenge in multiple regions globally^14^. These international comparisons reinforce the broader applicability of our results in demonstrating the importance of routine HBV screening and integrated treatment models in HIV care both within and beyond Africa.

Males showed significantly higher odds of HBV co-infection than females, a finding supported by prior studies^15,16^. Several factors may contribute to this disparity. First, studies have shown that men are more likely to engage in behaviors associated with HBV transmission than women; multiple sexual partnerships, inconsistent condom use, and injecting drug use^17^. Second, health-seeking patterns differ with women generally more likely than men to access healthcare services (that include antenatal care) and this often leads to earlier diagnosis and treatment in women^18^. Third, biological and immunological differences between males and females may contribute to disparities. Estrogen enhances immune responses, whereas testosterone has immunosuppressive effects^19^. Females are more likely to clear HBV after acute infection, whereas males are at greater risk of developing chronic HBV, which could explain the higher observed prevalence in men^20^. Taken together, these behavioral, healthcare access, and biological differences help explain the sex-based disparities in HBV prevalence among people living with HIV.

Individuals aged 25 years and older had significantly higher odds of co-infection. This likely reflects cumulative exposure over time, particularly in regions where HBV transmission occurs both perinatally and through horizontal means in adolescence and adulthood^21^. This trend has been consistently observed in prior studies from both Africa and other global regions. For example, Ondondo et al. (2024) in Kenya reported increasing HBV prevalence with age among PLWH, likely due to cumulative exposure and low childhood vaccination coverage prior to its inclusion in Kenya’s expanded program on immunization^22^. Similarly, Belyhun et al. (2016) noted a higher burden of HBV in older individuals across multiple African settings, reflecting horizontal transmission in adolescence and adulthood, especially in areas with late implementation of birth-dose HBV vaccination^5,11^. Global studies, including those by Thio et al. (2002), Shivakumar et al. (2024), and Tengan et al. (2017), further support this association between age and HBV co-infection risk among PLWH^10,14,21^. However, some studies have found less consistent age trends. For instance, Diop-Ndiaye et al. (2008) in Senegal and the EuroSIDA study (Price et al., 2013) did not identify a strong age-related pattern^12,13^. This may be due to different patterns of HBV transmission (e.g., more iatrogenic or MSM-associated in high-income countries), better vaccine uptake, or earlier screening and treatment in younger individuals. Despite some heterogeneity of findings from different studies, the overall evidence underscores the importance of targeting older adults for HBV screening and treatment in HIV programs, particularly in regions with historically low immunization coverage.

Our findings underscore the need to integrate HBV testing into routine HIV care. Historically, most standard ART regimens have included agents such as tenofovir disoproxil fumarate (TDF), which also suppress HBV replication^23,24^. As a result, HBV has often been managed incidentally in PLWH, without explicit diagnosis or monitoring. However, newer TDF-sparing regimens—such as long-acting injectables—lack HBV activity^25^. Initiating these regimens in individuals with undiagnosed chronic HBV may lead to viral reactivation and hepatitis flares due to the sudden withdrawal of HBV-suppressive agents^26^.

Our findings should be interpreted in the context of some limitations. HBV was determined by serological markers at a single time point, without differentiation between acute, chronic, or resolved infections. Participants were recruited from PEPFAR-funded clinical programs predominantly in urban settings, so our findings may not reflect the broader populations of people with and without HIV in each country.

HBV co-infection remains a major concern among PLWH in Sub-Saharan Africa, with variation by country, sex, and age. Scaling up HBV testing, vaccination, and integrated care is especially important in high-prevalence areas like Nigeria, where undiagnosed HBV could lead to reactivation when switching to TDF-sparing regimens. Future studies should assess long-term liver outcomes and the impact of newer ART regimens in co-infected populations.

## DECLARATIONS

### Ethics approval and consent to participate

The study was conducted in accordance with the ethical principles outlined in the Declaration of Helsinki. Ethical approval was obtained from Institutional Review Boards in each participating country: the Kenya Medical Research Institute Scientific and Ethics Review Unit (KEMRI SERU) in Kenya; the Uganda National Council for Science and Technology (UNCST) in Uganda; the National Health Research Ethics Committee of the National Institute for Medical Research (NIMR) in Tanzania; the National Health Research Ethics Committee (NHREC) in Nigeria; and the Walter Reed Army Institute of Research Institutional Review Board (WRAIR IRB) in the United States. Written informed consent was obtained from all participants prior to enrollment.

## ACKNOWLEDGEMENTS

We thank the study participants, local implementing partners, and hospital leadership at Kayunga District Hospital, Kericho District Hospital, AC Litein Mission Hospital, Kapkatet District Hospital, Tenwek Mission Hospital, Kapsabet District Hospital, Nandi Hills District Hospital, Kisumu West District Hospital, Mbeya Zonal Referral Hospital, Mbeya Regional Referral Hospital, Defence Headquarters Medical Center, and the 68^th^ Nigerian Army Reference Hospital.

We would also like to thank the AFRICOS Study Group – from the US Military HIV Research Program Headquarters Group^4,5^: Ajay Parikh, Brennan Cebula, Glenna Schluck, Jaclyn Hern, Jillian Chambers, Kara Lombardi, Kimberly Bohince, Leigh Anne Eller, Linsey Scheibler, Mary Schmitz, Michelle Imbach, Patricia Agaba, Sean Cavanaugh, Trevor Crowell; from the AFRICOS Uganda Group^12^: Agatha Mugagga Mukanza, Alfred Lutaaya, Anne Nakirijja, Benard Okanyakure, Betty Mwesigwa, Cate Kafeero, Christine Nabanoba, Christine Nanteza, Claire Nakazzi Bagenda, Estella Birabwa, Evelyn Najjuma, Ezra Musingye, Fred Magala, Freddie Ssentogo, Godfrey Zziwa, Grace Mirembe, Hannah Kibuuka, Harriet Nabirye, Hellen Birungi, Hilda Mutebe, Isaac Kato Kenoly, Jacqueline Namugabo, Michael Semwogerere, Michael Waiswa, Paul Wangiri, Phiona Namulondo, Prossy Naluyima, Richard Tumusiime, Ronald Ephraim Wasswa, Sylvia Namanda, Vamsi Vasireddy; from the AFRICOS South Rift Valley, Kenya Group^1,2^: Aaron Ngeno, Aggrey Koech, Alice Airo, Bornes Ngetich, Brayan Langat, Christopher Ochieng, Deborah Langat, Edwin Langat, Francis Opiyo, Grace Engoke, Ibrahim Daud, Ignatius Kiptoo, Irene Metet, Isaac Tsikhutsu, Janet Ngeno, Japhet Towett, Joan Kapkiai, Jonah Mwaswai, Joshua Rotich, Josphat Kosgei, Kennedy Labosso, Leonard Cheruiyot, Linner Rotich, Lucy Korir, Mary Leelgo, Mercy Chelimo, Michael Obonyo, Mike Bii, Raphael Langat, Rither Langat, Salome Ndungu, Samuel Kiprotich, Susan Ontango, Triza Rono, Wilfred Kirui, Zeddy Bett Kesi; from the AFRICOS Kisumu, Kenya Group^6,7^: Agnes Atieno, Celine Ogari, Charles Okwaro, Doris Njoroge, Elkanah Modi, George Suja, Iddah Aoko, Janet Oyieko, John Owuoth, Joseph Meyo, Kennedy Obambo, Lovet Nyawanda, Lucas Otieno, Michal Ohaga, Oscar Adimo, Paul Omolo, Solomon Otieno, Trizer Achieng, Valentine Sing’oei, Winnie Rehema; from the AFRICOS Mbeya, Tanzania Group^8,9^: Dorothy Mkondoo, Eliud Myegeta, Emmanuel Bahemana, Faraja Mbwayu, Gloria David, Goodluck Kisonga, Gwamaka Mwaisanga, Happy Laiton, Janeth Likiliwike, Jaquiline Mwamwaja, John Njegite, Johnisius Msigwa, Laban Mwandumbya, Lucas Maganga, Mtasi Mwaipopo, Naima Mkingule, Paschal Kiliba, Peter Edwin, Raphael Mkinga, Reginald Gervas, Restituta Minde, Rose Bruno, Vumilia Kaduma, Willyhelmina Olomi; from the AFRICOS Abuja, Nigeria Group^10,11^: Abdulwasiu Bolaji Tiamiyu, Aminu Suleiman, Blessing Edet Okon, Chisara Okolo, Felicia Anayochukwu Odo, Feyisayo Jegede, Helen Nwandu, Ifeanyi Okoye, Ijeoma Chigbu-Ukaegbu, Lawrence Umeji, Mfreke Asuquo, Ndubuisi Okeke, Onyinye Eze, Rosemary Akiga, Samirah Mohammed, Sunday Odeyemi, Victor Anyebe, Yakubu Adamu; from the AFRICOS Lagos, Nigeria Group: Abdulkadir Ramatu, Adewale Adelakun, Aire Commodore Edward Akinwale (Rtd), Blessing Irekpitan Wilson, Chiamaka Modesta Ibeanu, Concilia Uzoamaka Agbaim, Elekwa Chinenye Elizabeth, Igiri Faith, Jacinta Elemere, Jumoke Titilayo Nwalozie, Lt Col Sulaimon Awesu, Maj Christian Etim Efut, Ndubuisi Rosemary Obiageli, Nkechinyere Harrison, Nnadi Theodora Nkiru, Obende Theresa Owanza, Obilor Ifeoma Lauretta, Olutunde Ademola Adegbite, Rachael Eluwa, Uzoegwu Amaka Natalie, Victoria Idi, Yakubu Adamu, Zahra Parker.

Full institutional affiliations correspond to the superscript numbering in the main author list.

## Disclaimer

This material has been reviewed by the Walter Reed Army Institute of Research. There is no objection to its presentation and/or publication. The opinions or assertions contained herein are the private views of the author, and are not to be construed as official, or as reflecting true views of the Department of the Army or the Department of Defense. The investigators have adhered to the policies for protection of human research participants as prescribed in AR 70–25.

## Prior Presentation

This work was previously presented, in part, at the Conference on Retroviruses and Opportunistic Infections in Denver, Colorado, 3-6 March 2024.

## Data Availability Statement

The datasets generated and/or analyzed during the current study are available from the corresponding author on reasonable request. To request a minimal data set, please contact the Data Coordinating and Analysis Center (DCAC) at and indicate the AFRICOS (RV329) study along with the name of the manuscript.

## Clinical trial number

not applicable

## ABBREVIATIONS

AFRICOS: African Cohort Study
ALT: Alanine aminotransferase
ART: Antiretroviral therapy
AST: Aspartate aminotransferase
CI: Confidence interval
DILI: Drug-induced liver injury
ELISA: Enzyme-linked immunosorbent assay
HBsAg: Hepatitis B surface antigen
HBV: Hepatitis B virus
HIV: Human immunodeficiency virus
IQR: Interquartile range
IRB: Institutional Review Board
KEMRI: Kenya Medical Research Institute
NIMR: National Institute for Medical Research
NHREC: National Health Research Ethics Committee
OR: Odds ratio
PEPFAR: President’s Emergency Plan for AIDS Relief
PLWH: People living with HIV
RR: Relative risk
TDF: Tenofovir disoproxil fumarate
ULN: Upper limit of normal
UNCST: Uganda National Council for Science and Technology
WRAIR: Walter Reed Army Institute of Research

